# Intrahost evolution of the HIV-2 capsid correlates with progression to AIDS

**DOI:** 10.1101/2021.12.28.21268379

**Authors:** M. T. Boswell, J. Nazziwa, K. Kuroki, A. Palm, S. Karlson, F. Månsson, A. Biague, Z. J. da Silva, C.O. Onyango, T.I. de Silva, A. Jaye, H. Norrgren, P. Medstrand, M. Jansson, K. Maenaka, S. L. Rowland-Jones, J. Esbjörnsson, the SWEGUB CORE group

## Abstract

**Background:** HIV-2 infection will progress to AIDS in most patients without treatment, albeit at approximately half the rate of HIV-1 infection. HIV-2 p26 amino acid variations are associated with lower viral loads and enhanced processing of T cell epitopes, which may lead to protective Gag-specific CTL responses common in slower disease progressors. Lower virus evolutionary rates, and positive selection on conserved residues in HIV-2 *env* have been associated with slower progression to AIDS. We therefore aimed to determine if intrahost evolution of HIV-2 *p26* is associated with disease progression.

**Methods:** Twelve treatment-naïve, HIV-2 mono-infected participants from the Guinea-Bissau Police cohort with longitudinal CD4+ T cell data and clinical follow-up were included in the analysis. CD4% change over time was analysed via linear regression models to stratify participants into relative faster and slower disease progressor groups. *Gag* amplicons of 735 nucleotides which spanned the *p26* region were amplified by PCR and sequenced. We analysed *p26* sequence diversity evolution, measured site-specific selection pressures and evolutionary rates, and determined if these evolutionary parameters were associated with progression status. Amino acid polymorphisms were mapped to existing p26 protein structures.

**Results:** In total, 369 heterochronous HIV-2 *p26* sequences from 12 male patients with a median age of 30 (IQR: 28–37) years at enrolment were analysed. Faster progressors had lower CD4% and faster CD4% decline rates. Median pairwise sequence diversity was higher in faster progressors (5.7×10^−3^ versus 1.4×10^−3^ base substitutions per site, *P*<0.001). *p26* evolved under negative selection in both groups (dN/dS=0.12). Virus evolutionary rates were higher in faster than slower progressors – synonymous rates: 4.6×10^−3^ vs. 2.3×10^−3^; and nonsynonymous rates: 6.9×10^−4^ vs. 2.7×10^−4^ substitutions/site/year, respectively. Virus evolutionary rates correlated negatively with CD4% change rates (ρ = -0.8, *P*=0.02), but not CD4% level. However, Bayes factor (BF) testing indicated that the association between evolutionary rates and CD4% kinetics was supported by weak evidence (BF=0.5). The signature amino acid at p26 positions 6, 12 and 119 differed between faster (6A, 12I, 119A) and slower (6G, 12V, 119P) progressors. These amino acid positions clustered near to the TRIM5α/p26 hexamer interface surface.

**Conclusions:** Faster *p26* evolutionary rates were associated with faster progression to AIDS and were mostly driven by synonymous substitutions. Nonsynonymous evolutionary rates were an order of magnitude lower than synonymous rates, with limited amino acid sequence evolution over time within hosts. These results indicate the HIV-2 p26 may be an attractive vaccine or therapeutic target.

## Introduction

HIV-1 and HIV-2 are retroviruses which transmitted from non-human primates to humans in the 20th century. HIV-1 has four main groups, of which M accounts for 98% of global infections. HIV-1 group M descended from SIVcpz which circulates in chimpanzees (1). HIV-2 is descended from SIVsm which circulates in sooty mangabeys, and has nine groups (A – I), of which A and B account almost all human infections (1). HIV-1 has caused a global pandemic, whereas HIV-2 has remained an endemic infectious disease in West Africa, with limited spread outside the region (2). In addition to stark differences in transmissibility, the two viruses differ in disease progression rates (3,4). In the absence of treatment, HIV-1 will progress to AIDS twice as fast as HIV-2. However, HIV-2 disease progression is highly variable, with some patients developing AIDS in similar timeframes as HIV-1 and other progressing far more slowly.

After infecting a host, these viruses rapidly diversify and circumvent host immune responses, founding numerous sub-populations known as the virus quasispecies (5). An extensive body of literature has connected intrahost evolution of HIV-1 to disease progression and phenotypic trait development (6–12). Synonymous evolutionary rates (nucleotide substitutions which do not change amino acid sequences) in HIV-1 *env* correlate with time to AIDS, CD4+ count decline rates and viral load, while nonsynonymous evolutionary rates are not associated with disease progression. Similar associations have been reported for intrahost evolution HIV-1 *gag* (13). In HIV-1 infection, higher replicative capacity of founder viruses explains much of the variation in disease progression rates, and is strongly associated with increasing T cell activation and exhaustion (14). Together these results suggest that faster virus replication drives HIV-1 disease progression due to heightened immune activation, rather than evolution being driven by immune escape (11).

HIV-2 *env* evolutionary rates in faster disease progressors are approximately double that of slower progressors (15). In HIV-2 infection the relationship between immune activation and progressive CD4+ T cell loss is similar to that in HIV-1, at equivalent degrees of immune suppression (16). This suggests that evolutionary rates, immune activation, and subsequent disease progression are similarly linked in HIV-1 and HIV-2 infection.

The HIV-1 capsid is a fullerene cone structure made up of 216 p24 hexamers, and 12 p24 pentamers (17). The capsid is critical in the viral replication cycle, with several indispensable functions including nucleotide supply to the replicating virus, nuclear import to facilitate integration, cyclophilin-A (CyPA) binding, and immune evasion (18– 21). HIV-1 and HIV-2 capsids bind to the innate sensor NONO in the nucleus, which triggers the cGAS-STING pathway to activate innate immune responses in macrophages and dendritic cells (21). HIV-2 binds NONO with higher affinity than HIV-1. HIV-2 capsids are also more sensitive to restriction factor TRIM5α than HIV-1 (22). Polymorphisms in HIV-2 p26 have been linked to disease progression. Specifically, participants with low viral loads often have prolines in p26 positions 119, 159 and 178 (23). Prolines at position 159 and 178 are associated with enhanced proteasomal processing of CD8^+^ T cell epitopes and greater Gag-specific T cell responses, and is a frequent target for T cell responses which are associated with low viral loads (24). Additionally, proline at position 119 in p26 alters the conformation of the capsid structure and is associated with increased sensitivity to TRIM5α (25,26).

We tested this hypothesis by analysing the following in relation to HIV-2 disease progression: i) pairwise sequence diversity evolution; ii) site-specific selection pressures; iii) synonymous and nonsynonymous evolutionary rates in *p26* and iv) p26 amino acid sequence variation in the virus quasispecies.

## Methods

This section provides a summary of the main laboratory and statistical methods used in this study. Further details are available in the supplementary materials.

### Study participants

Study participants were recruited to the Guinea-Bissau Police cohort between 1990-2009. Informed consent was obtained from the participants and the study was approved by the research ethic committees of the Ministry of Health in Guinea-Bissau, Lund University and the Karolinska Institute, Sweden. Twelve HIV-2 infected participants were included in this analysis based on availability of plasma samples. Selection criteria included that the participants were: HIV-2 monoinfected, antiretroviral therapy (ART) naïve at the time of plasma sample collection and had longitudinal CD4+ T cell measurements available which allowed for estimation of disease progression (**Table S1**). Clinical and immunological staging of HIV disease was done according to the WHO criteria (27). A total of seven participants had an estimated date of HIV-2 seroconversion (**seroincident**) and five participants were HIV-2 positive at enrolment (**seroprevalent**). T cell data was analysed from the first time point after HIV-2 detection, in seroincident participants this time point was the first one after their documented seroconversion, and in seroprevalent participants at enrolment into the cohort.

### Analysis of disease progression markers in HIV-2

Both absolute CD4+ T cell counts and CD4+ T cell percentage (CD4%) are reliable immunological markers of HIV disease progression. In resource-limited settings, CD4+ T cell percentages (CD4%) are less sensitive to specimen handling, patient age, or time of sampling when compared to absolute CD4 counts (28–31). We therefore chose to analyse disease progression using CD4% change over time. Briefly, participants’ longitudinal CD4% were analysed in per-participant linear regression models. The CD4% level at the midpoint in follow-up time after HIV-2 detection and the CD4% change rate (slope of the regression line) were extracted from these models and analysed as markers of disease progression.

To create disease progression groups, participants were ranked and classified according to three approaches as previously described (15) i) from highest midpoint CD4% to the lowest (those above the mean were classified as slower progressors and those below the mean as faster progressors); ii) from highest positive change rate to the lowest negative change rate; iii) the midpoint CD4%, and CD4% change rate were then transformed into proportional values, added together, and averaged for each participant. This gave a combined coefficient for each participant which was weighted equally according to midpoint CD4% and the rate of change which accounts for differences in disease stage at enrolment. The combined coefficient was used to rank participants and stratify them into relative progression with distinct disease phenotypes – faster and slower disease progression (**Table S2**). All analyses which refer to progression groups used this combined coefficient for stratification.

### RNA extraction and PCR amplification

Briefly – plasma samples had previously been collected from participants and stored at -80°C; we extracted RNA using the RNeasy Lipid Tissue Mini Kit (Qiagen, Venlo, Netherlands) with minor modifications to the manufacturer’s instructions. Following RNA extraction, a nested PCR was performed with 5µL of extracted RNA. The first step involved a one-step reverse transcription PCR (RT-PCR) using the SuperScript IV One-Step RT-PCR System with Platinum Taq DNA Polymerase (supplementary materials [**Tables S3-6**]). Immediately following the RT-PCR a second nested reaction using an inner set of HIV-2 *p26* primers was performed using the Dream Taq PCR kit (Thermo Fisher Scientific, Waltham, MA (**Figure S1**). Primer sequences are listed in Table S4.

### Cloning and sequencing

Amplicons of 846 nucleotides from position 1411–2257 on HIV-2 BEN.M30502 were cloned into pCR^®^4 TOPO^®^ vector using TOPO-TA cloning kit (Invitrogen, Carlsbad, CA, USA); 23 white colonies were randomly picked and amplified by colony-PCR using the Advantage 2 PCR kit (Takara, Kusatsu, Japan) (**Figure S2**), to confirm presence of the insert. The plasmids containing inserts were sequenced by Sanger sequencing using the inner PCR primers (**Table S4**) (Macrogen Europe, Amsterdam, Netherlands).

### Sequence analysis

Raw sequence data were analysed in Geneious Prime v. 2019.2 (Geneious, Biomatters, Auckland, New Zealand), mapped to the HIV-2 BEN.M30502 reference sequence and trimmed for quality (32). Contigs were constructed for each participant from single forward and reverse reads. Poor quality reads and contigs with high quality (HQ) scores of less than 80% were not included in further downstream analysis. This score indicates that 80% of bases are high quality and that the likelihood of a false positive base reading is 1:10 000 (33). Mixed peaks in electropherograms were resolved in favour of the consensus nucleotide at the base. Contigs were assembled into participant-specific alignments and mapped to HIV-2 BEN.M30502 to ensure that all participant sequences mapped to the same coding region of *gag*. Recombination between sequences grouped in an alignment can violate the assumptions made in phylogenetic analysis (34,35). We therefore screened each participant-specific dataset for recombination in RDP4 using a combination of the following methods: RDP, GENECONV, BootScan, MaxChi and Chimaera (35–40). All sequences showing evidence of recombination were removed from further analysis.

### Bayesian phylogenetic analysis

All Bayesian phylogenetic parameters were specified in BEAUti v. 1.10.4 and run in BEAST v1.10.4 with parallel processing by BEAGLE (41,42). Model output logs were analysed in Tracer v1.7 (43) and assessed for convergence by visual inspection of the posterior distribution chains and if effective sample sizes (ESSs) were >100, after 10% burn-in. All analyses were run in duplicate to assess reproducible convergence. Bayesian phylogenetic models were used to **(i)** perform viral subtyping **(ii)** reconstruct the most recent common ancestor (MRCA) for the participants’ combined sequence alignment, **(iii)** measure pairwise sequence diversity within patient sequence alignments **(iv)** estimate site-specific dN/dS ratios in p26 for each participant **(v)** estimate relaxed molecular clock estimates for individual participants and **(vi)** estimate participant-specific molecular clock rates via a hierarchical phylogenetic model (HPM)

i. Viral subtyping was done by sampling HIV-2 *gag* sequences from the clonal sequences. *Gag* sequences for HIV-2 groups A – G as well as SIVsmm and HIV-1 were downloaded from the Los Alamos National Laboratory (LANL) sequence database, and an alignment of the participant HIV-2 sequences with these reference sequences was created (**Table S7**). Viral subtyping was performed using a Bayesian phylogenetic approach. A strict molecular clock model, HKY substitution model and constant population size were specified. The Markov chain was run for 2×10^6^ iterations.
ii. The following parameters were used for the MRCA reconstruction -strict molecular clock model, GTR nucleotide substitution model, no site heterogeneity or codon partition, constant population size. The Markov chain was run for 2.5×10^6^ iterations. The MRCA was used as a reference sequence for variant identification and mapping of amino acid variants over time (discussed below).
iii. To measure pairwise sequence diversity the sequences from the patient samples are labelled according to the time point of collection, these are then aligned using CLUSTAL-W. Using GARLI V2.01, 200 maximum likelihood bootstrap trees are generated for each dataset. The diversity estimates in base substitution per base site are obtained from each of the 200 trees using the BIOTREE:IO function of the BioPerl package. These estimates are then summarised in R to get the mean diversity and the 95% confidence intervals.
iv. To estimate selection pressure the dN/dS ratio is calculated by dividing the nonsynonymous evolutionary (dN) rate by the synonymous (dS) evolutionary rate (a ratio of less than 1.0 will indicate negative selection; a ratio equal to 1.0 neutral selection, and a ratio greater than 1.0 positive selection at a site). We measured site-specific selection pressures using Renaissance counting procedures performed with the following model parameters – a strict molecular clock model, GTR nucleotide substitution model, no site heterogeneity, a 1,2,3 codon partition, constant population size and 4×10^7^ MCMC chains (44).
v. To determine combined, synonymous and nonsynonymous evolutionary rates derived from the participant-specific relaxed molecular clock models we used software packages developed by Lemey *et al*. (11). Briefly, for each participant nucleotide alignment a relaxed molecular clock model was created in BEUti and run in BEAST. Model parameters included an HKY nucleotide substitution model, gamma site heterogeneity, a constant population size and 2×10^8^ MCMC chains. For each participant 10 000 trees were simulated as a posterior sample distribution. From these 10 000 trees 200 were selected randomly after a 10% burn-in and separated into nucleotide substitution unit denoted and time unit denoted trees. The substitution trees were then separated further using HyPhy into expected synonymous and nonsynonymous substitution trees. Next, these substitution trees were analysed separately in conjunction with their respective time unit trees to generate estimates of the combined, synonymous, and nonsynonymous evolutionary rates. Lastly, the substitution trees were used to estimate divergence as a function of time from the first time point to the last time point in the alignment. This allowed us to plot divergence over time.
vi. A significant limitation of the relaxed molecular clock estimates described above are interparticipant variability in number of sequences, number of time points, and total follow-up time. The HPM allows for feedback across participant average estimates to improve participant specific estimates, thereby making use of a larger data set (all sequences linked in the HPM) to inform smaller partitions in the data (participant specific estimates). Another main difference in the HPM estimates versus those above are that HPMs assume a strict molecular clock per partition which does not vary for each taxon while the previous estimates assumed an uncorrelated relaxed molecular clock per participant. We partitioned the HPM by using fixed factor terms for the combined coefficient stratification, as well as the log values of the midpoint CD4% and CD4% change rate. Model parameters included a strict molecular clock model, HKY nucleotide substitution model, gamma site heterogeneity, a 1,2 and 3 codon partition, constant population size. The Markov chain was run for 6×10^8^ iterations. We calculated Bayes Factors to test whether the midpoint CD4% and CD4% change rate were significant explanatory variables in evolutionary rate estimates. We repeated the HPM analysis with a narrower range of prior values which could be assessed by the model. Our initial HPM used a hyperprior scale parameter of 1000, and we subsequently adjusted these to scales of 100 and 10 – this allows for a narrower distribution of values to be assessed for prior parameters (45).

### Amino acid sequence analysis

We translated and aligned all *p26* sequences to the MRCA to identify single amino acid polymorphisms (SAAPs). We categorised SAAPs as majority variants if they occurred in more than 50% of sequences; and minority variants if the prevalence was below 50%. Polymorphisms found in a single sequence were designated as private variants. Private single nucleotide polymorphisms identified in virus quasispecies using cloning and Sanger sequencing are often not found using next-generation sequencing platforms and may represent a combination of PCR and sequencing errors (46).

To test whether the frequency of variants significantly differed between progression groups, we divided the sequence alignment by progression status (faster vs. slower progressors). We then created new alignments of 1000 sequences per progression group (randomly selected via bootstrap sampling with replacement). We then compared the amino acid frequencies by progressor group using the Viral Epidemiology Signature Pattern Analysis (VESPA) tool (47). VESPA identifies signature patterns, which differ between alignments and reports frequencies of the specific amino acids in each alignment. HIV-2 p26 structural models were generated in Pymol v. 1.8, using protein sequence PDB ID: 2WLV (48). Residues which have been linked to important functional regions/structures on the HIV-1 p24 protein were used to infer sites on HIV-2 p26 which may have a similar role. This was done by aligning reference sequences HIV-1.NL4-3 and HIV-2 BEN.M30502.

### Caio cohort sequence analysis

We investigated whether the signature amino acid variants identified by the VESPA analysis were associated with disease progression markers in a larger, external cohort. This was done by testing the association of these p26 amino acid variants with CD4% and HIV-2 plasma viral loads in 86 HIV-2 positive participants from the Caio cohort (23,49).

### Statistical analysis

All statistical analyses were 2-sided, and data visualisations were done in R studio v. 4.0.3, unless specified otherwise (50,51). Baseline characteristics of participants were summarised as means and standard deviations, medians, and interquartile ranges, and counts with percentages. Pairwise testing was performed using the Mann Whitney U (MW) test for non-normally distributed data, and the Student’s T test for normally distributed data. To compare distributions of repeated measurements between the progression groups we used Friedman tests with effect sizes reported by Kendall’s W value (small effect: W = 0.1-0.3; moderate effect: 0.3-0.5 and large effect >0.5). The frequency of categorical variables was assessed by chi squared tests. Proportional differences were also assessed using the Fisher’s Exact Test (FET) and reported as odds ratios (ORs). Correlation statistics were calculated using the Pearson (parametric) or Spearman’s (non-parametric) correlation coefficients dependent on the variables’ distribution. Pairwise sequence diversity evolution was quantified using linear mixed effects models with model fit assessed by the likelihood ratio test (LRT). A False discovery rate (FDR) was used to correct for multiple comparisons. Statistical significance was determined as *P*<0.05.

## Results

Our analysis included 12 HIV-2 infected participants from the Guinea-Bissau Police cohort. All participants were male with a median age at enrolment of 30 (IQR: 28–37) years. The cohort median midpoint CD4% was 27.5%, and the median CD4% change rate was -0.05% per year. After calculation of the combined coefficient and assignment to progression groups there were six faster progressors and six slower progressors. Faster progressors had a significantly lower midpoint CD4% than slower progressors (20.7% vs. 30.6%, *P*=0.02, MW). In addition, the faster progressors’ CD4% decreased by 1.4% per year, whereas the slower progressors’ CD4% increased by 0.6% per year (*P*=0.02, MW)). One of the slower progressors developed severe immunosuppression (CD4% below 15), whereas 5/6 faster progressors reached a CD4% below 15% during follow up (**Figure 1**). In seroincident participants the median time from HIV-2 detection to the first sample was 4.5 (IQR: 3.9–6.7) years, and in seroprevalent participants the median time from HIV-2 detection (enrolment) to sample was 14.4 (IQR: 2.4–15.6) years. The mean follow-up time from first to last sequencing sample was 5.9 years, 6.1 years for faster progressors and 5.7 for slower progressors.

**Figure 1.**
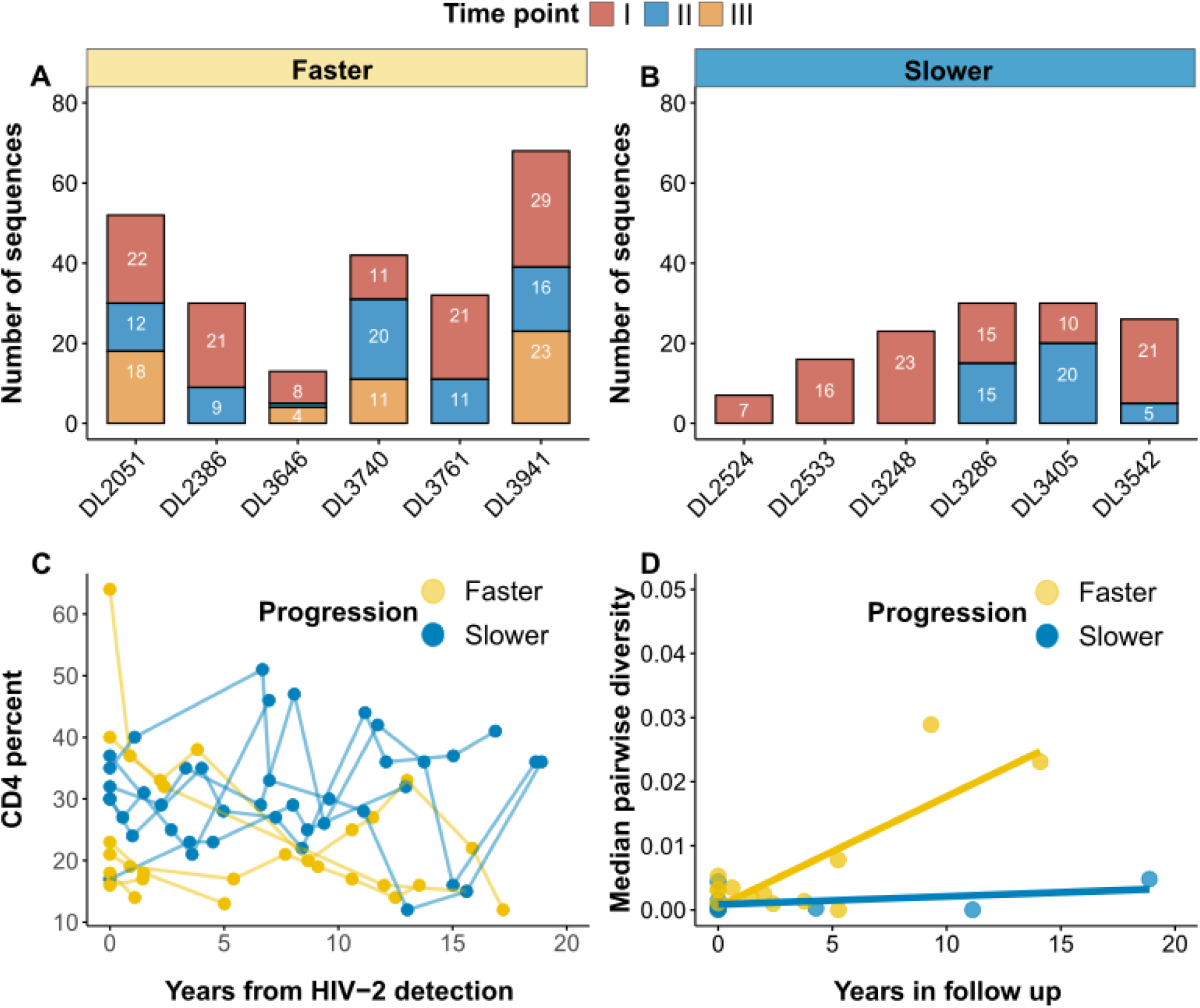
Summary of number of sequences, CD4% kinetics, and pairwise diversity by progression status. **A - B)** Bar plots summarise the number of sequences per patient and time point. Panels are stratified by progression status as determined by the combined coefficient. There are six faster and slower progressors. Slower progressors had fewer sequences (numbers inside the bars) and time points for analysis than faster progressors. **C)** CD4% kinetics shown for faster and slower progressors as defined by the combined coefficient. Faster progressors had a significantly lower CD4% and faster CD4% decline rates than slower progressors (*P<*0.05, MW). **D)** Scatter plots with fitted linear regression lines showing pairwise diversity increasing over time, presented as nucleotide substitutions per site (y-axis) and time in years between samples (x-axis). Pairwise sequence diversity was higher than slower progressors (*P*<0.05, MW).

After aligning the sequences and trimming for quality, the analysed *gag* sequences spanned 735 nucleotides in a single open reading frame which mapped to nucleotide 1460-2194 on HIV-2 BEN.M30502. These sequences are available from Genbank with accession numbers OL872372-872739 and OM146012. This sequence included the first 687 nucleotides, from 5’ to 3’, of the p26 region of *gag*. Twenty-five plasma samples yielded PCR products from the 12 participants; with PCR products from longitudinal sample time points generated from nine participants (**Table 1**). In total 575 clones were generated and sequenced, whereof 173 were removed due to poor sequence quality or presence of stop codons. Of the remaining 402 sequences, 33 were possible recombinant sequences, and were removed from further analysis. This resulted in a final dataset of 369 sequences from 12 study participants (median: 30 sequences/participant; **Figure 1**). Participant sequences formed monophyletic clusters with high posterior support values, suggesting that contamination or labelling errors did not occur during sample handling. No clear clustering pattern by progression status was observed (**Figure S3**). Moreover, the subtype analysis indicated that all sequences clustered with HIV-2 group A reference sequences (**Table S7**) (**Figure S4**).

**Table 1:** Summary results for the study participants.

| Study ID | HIV-2 serostatus <sup>1</sup> | Midpoint CD4% <sup>2</sup> | CD4% change rate <sup>3</sup> | Progression <sup>4</sup> |
| --- | --- | --- | --- | --- |
| DL2051 | Prevalent | 30.7 | -1.8 | Faster |
| DL2386 <sup>5</sup> | Incident | 25.3 | -1.6 | Faster |
| DL2524 | Prevalent | 32.7 | 0.7 | Slower |
| DL2533 | Prevalent | 25.1 | 1.1 | Slower |
| DL3248 | Incident | 29.4 | 1 | Slower |
| DL3286 | Incident | 32.7 | -0.1 | Slower |
| DL3405 | Incident | 33.1 | 1.5 | Slower |
| DL3542 | Prevalent | 30.8 | -1 | Slower |
| DL3646 | Prevalent | 18.2 | -0.3 | Faster |
| DL3740 | Incident | 16.5 | 0.7 | Faster |
| DL3761 | Incident | 16 | -3.7 | Faster |
| DL3941 | Incident | 17.4 | -1.9 | Faster |

<sup>1</sup> HIV-2 serostatus includes seroincident (where an estimated date of seroconversion is available), and seroprevalent participants (where a participant was HIV-2 positive at time of enrolment). <sup>2</sup> Midpoint CD4% calculated from per-patient regression models. <sup>3</sup> CD4% change rate per year is the linear regression coefficient of CD4% over time. <sup>4</sup> Progression status as determined by the combined coefficient <sup>5</sup> Sample I for participant DL2386 did not have a time stamp. We therefore used the midpoint in follow-up time for this participant with an uncertainty correction of $\pm 9$ years in all phylogenetic models which required a date for the sample.

### Pairwise sequence diversity is associated with disease progression status

Sequence diversity increased over time (Spearman correlation: ρ = 0. 26, *P*<0.001, **Figure 1**). The pooled median sequence diversity was significantly higher among faster than slower progressors (5.7×10^−3^ versus 1.4×10^−3^ base substitutions per site, MW *P*<0.001). Analysing linear regression models of diversity over time, the average sequence diversity increased by 1.7×10^−3^ (95% CI: 1.7-1.8×10^−3^, *P*<0.001) substitutions per site, per year (s/s/y), for the fasters progressors, and increased by 1.3×10^−4^ s/s/y (95% CI: 1.1-1.4×10^−4^, *P*<0.001) for the slower progressors. To account for the unequal contribution of sequences, time points, and inter-participant variability, we analysed mixed effects models. A random slope and intercept model (time (i.e., slope of diversity) allowed to vary by participant (random effect)) provided the best model fit (*P*<0.001, LRT). In this model, progression status was not associated with a significant effect on diversity change over time (faster progression diversity increased by 1.2×10^−3^ relative to slower progression, 95% CI = -9.2×10^−4^-3.5×10^−3^, *P*=0.2).

### Progression status is associated with synonymous and nonsynonymous evolutionary rates

The results for the relaxed clock estimates are reported for nine participants (six faster and three slower) who had sequences from multiple time points available. The median evolutionary rate in *p26* for all participants was 4.0×10^−3^ (IQR: 2.6-6.9×10^−3^) s/s/y. Evolutionary rates correlated negatively with CD4% change rates, but not midpoint CD4% (Spearman correlation: ρ = -0.8 and -0.5 respectively, *P*=0.02 and 0.17). Faster progressors had a significantly higher evolutionary rate than slower progressors (5.4×10^−3^ vs. 2.5×10^−3^ s/s/y, W = 0.4, FT *P*<0.001, **Figure 2**). The median synonymous evolutionary rate for all participants was 3.5×10^−3^ (IQR: 2.3–6.1 ×10^−3^) s/s/y and was also significantly higher in faster than slower progressors (4.6×10^−3^ vs. 2.3×10^−3^ s/s/y, W = 0.4, FT *P*<0.001, **Figure 2**). The median nonsynonymous evolutionary rate for all participants was 4.1×10^−4^ (IQR: 2.4–8.8×10^−4^) s/s/y and was significantly higher in faster than slower progressors (6.9×10^−4^ vs. 2.7×10^−4^ s/s/y, W = 0.4, FT *P*<0.001, **Figure 2**). The dN/dS ratio for *p26* in was 0.12 (IQR: 0.08–0.21), and the difference between faster and slower progressors was small but significant (0.13 vs. 0.11, W = 0.2, FT *P*<0.001). Relaxed evolutionary rate estimates are summarised for each participant in the supplementary materials (**Table S8**). The effect sizes for progression groups on evolutionary rate comparisons were moderate (W = 0.4), and for dN/dS ratios small (W = 0.2). Synonymous and nonsynonymous divergence in *p26* increased linearly with time in both faster and slower progressors (**Figure 2**). Linear regression models were fitted to each participant’s divergence over time (r^2^ for synonymous and nonsynonymous divergence was 0.94 and 0.82, respectively).

**Figure 2.**
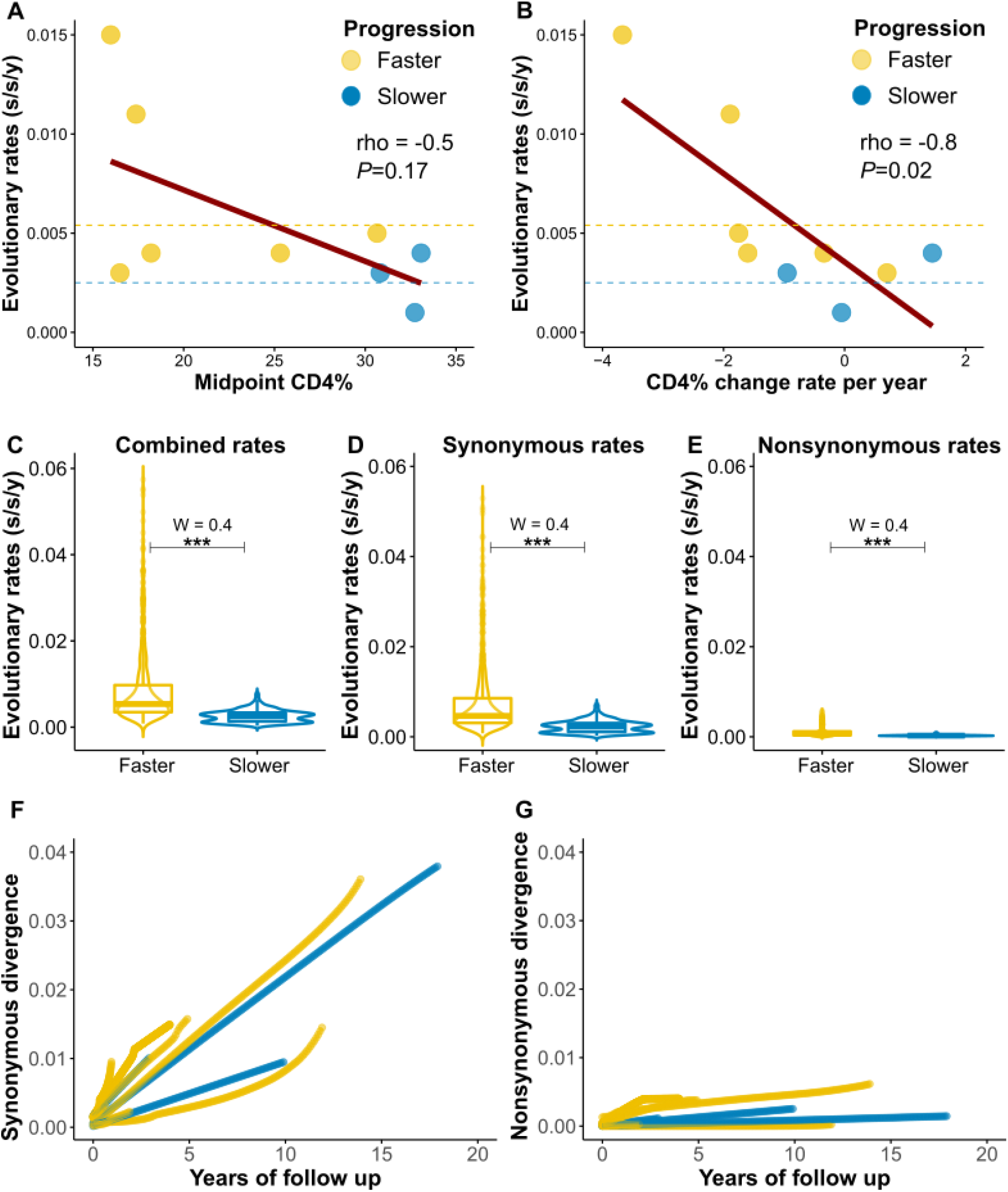
Evolutionary rates in relation to disease progression markers. Results are shown for the nine participants with multiple sequence time points **A - B)** Median relaxed clock evolutionary rates (y-axis) correlated significantly with CD4% change rate, but not CD4% (x-axes). Dashed lines show the median evolutionary rates for faster and slower progressor groups. **C - E)** Violin plots showing evolutionary rate distributions from 200 randomly sampled trees per patient (y-axis) by their respective progression group (x-axis). Combined, synonymous and nonsynonymous evolutionary rates were significantly higher in faster progressors (FT *P*<0.001). **F - G)** Line plots of synonymous and nonsynonymous divergence (measured in substitutions per site) over time by progressor group. The accumulated divergence (y-axis) from the first analysed sample for each participant indicated by time = 0 on the x-axis. Faster progressors are coloured in yellow, slower progressors in blue. *** = *P*<0.001.

We next analysed a series of HPMs to generate estimates of evolutionary rates in *p26* for each participant with multiple time points and tested the correlation between evolutionary rates, midpoint CD4% and CD4% change rate. The median HPM evolutionary rate for all participants was 3.1×10^−3^ s/s/y (95% HPD: 1.9-4.5×10^−3^) and correlated well with the relaxed clock estimates (Spearman correlation: ρ = 0.8, *P*=0.008). We analysed the midpoint CD4%, CD4% change rates and the combined coefficient stratification as fixed effects to determine if the disease progression markers explained variation in evolutionary rates. There was weak evidence that disease progression markers were associated with evolutionary rate variation. The Bayes Factor for midpoint CD4%, CD4% change rate and combined coefficient stratifications ranged from 0.4 to 0.5. Median HPM evolutionary rates did not correlate with the midpoint CD4% or CD4% change rate (spearman correlation: ρ = -0.3 and -0.7 respectively, *P*>0.05). The median evolutionary rate estimates were similar across hyperprior scale values. The median clock rate across all participants for a scale of 10, 100 and 1000 was 2.9×10^−3^ (95% HPD: 1.6–4.9×10^−3^), 3.1×10^−3^ (95% HPD: 1.8– 4.9×10^−3^) and 3.1×10^−3^ (95% HPD: 1.9–4.5×10^−3^) s/s/y, respectively.

### Estimation of site-specific selection pressures in *p26*

Next, we used Renaissance counting procedures to quantify selection pressures in *p26*, per participant, by estimating site-specific dN/dS ratios (44). We included participants with two or more time points for this analysis (n = 9). Negative selection predominated for all participants, with only one faster progressor showing a signature of positive selection at one site (DL2051 at position five).

### HIV-2 p26 amino acids signatures differ by progression status

All sequences were used to reconstruct the MRCA sequence at the root of the maximum clade credibility tree (**Figure S3**). The MRCA sequence aligned well with HIV-2 BEN.M30502, differing at 10 amino acid positions, and was used for amino acid variant identification. In total, 61 positions among the participants’ amino acid sequences differed from the MRCA sequence. After excluding private variants (found in only one sequence), faster progressor amino acid sequences were more likely to differ from the MRCA than slower progressor sequences (35 vs. 19 positions differed, OR = 2.0, 95% CI = 1.1–3.8, *P*=0.03).

Sixty-nine unique amino acid variants were identified, and 28 of these were private variants. Of the 41 remaining variants, two were major variants (present in more than 50% of all sequences) and 39 were minor variants (present in less than 50% of all sequences) Two positions on p26, 85 and 96, in the CyPA binding loop showed variation from the MRCA (**Figure 3**). Based on the results from the VESPA analysis, the amino acid at three positions differed between faster and slower disease progressors. At positions 6, 12 and 119, in slower progressors the most common amino acids were glycine, valine, and proline, and in faster progressors the most common were alanine, isoleucine, and alanine (**Figure 4**). These amino acid positions localised to the N-terminal domain of HIV-2 p26 (**Figure 4**)(52). In addition, position 6 and 12 are located at the p26 hexamer interface surface, while 119 is next to the CyPA binding loop (52–54). Many of the variants were stable in follow-up and there was little evidence of specific variants being consistently selected for - agreeing with the Renaissance counting results (**Figure 3**).

**Figure 3.**
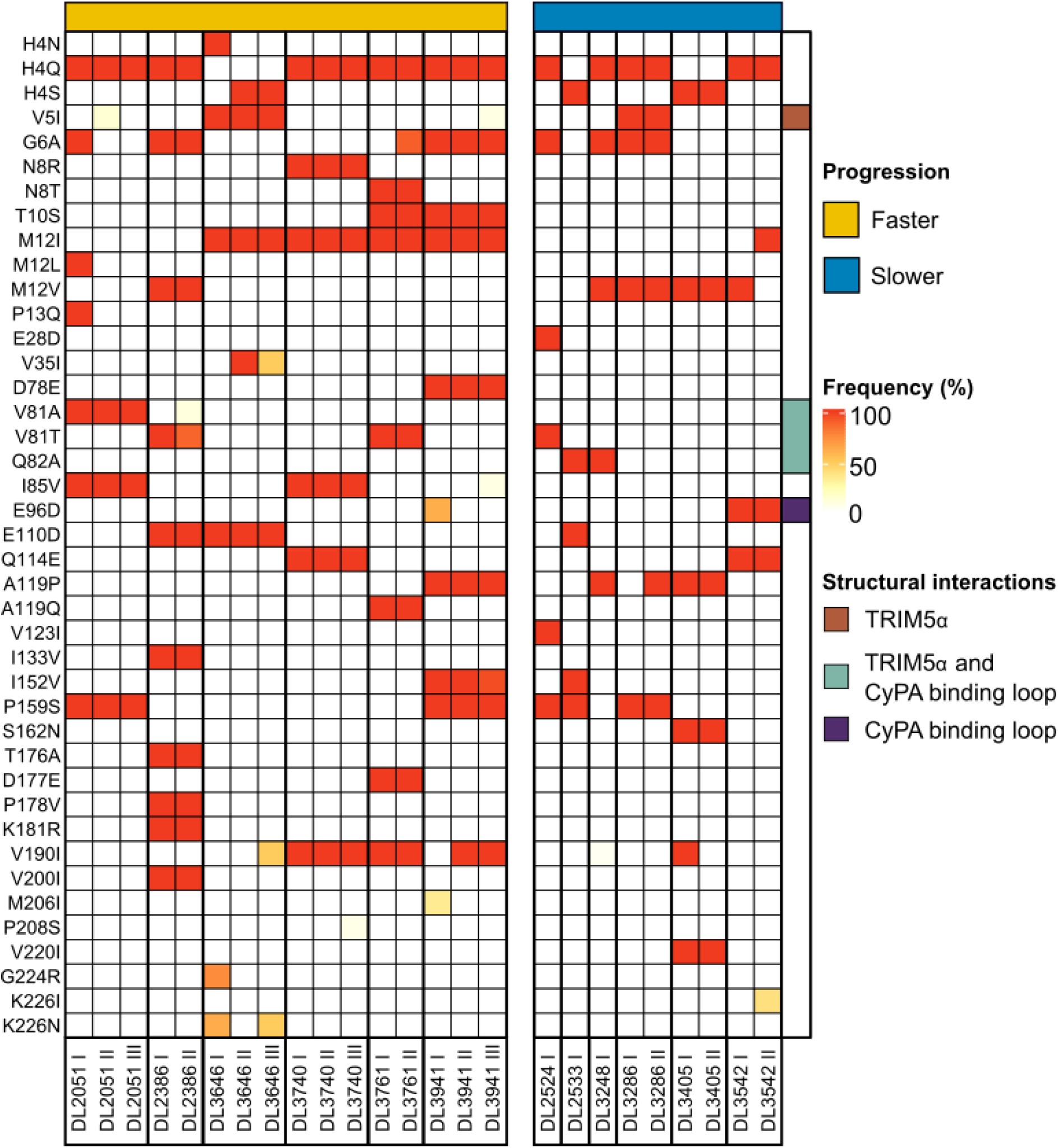
Heatmap of HIV-2 p26 amino acid variant frequencies over time. The y-axis shows the amino acid variants’ frequencies, with the MRCA amino acid sequence used as a reference for selected positions. Each column is a participant-specific time point. Participants with their respective time points (I – III) are shown on the x-axis, split by progression status. Variant frequencies are shown as a percentage of the sequenced quasispecies, with the scale on the right. Progressor groups are annotated on the top x-axis, yellow = faster progressors, blue = slower progressors. p26 amino acids which directly bind to TRIM5α and CyPA are indicated as well.

**Figure 4.**
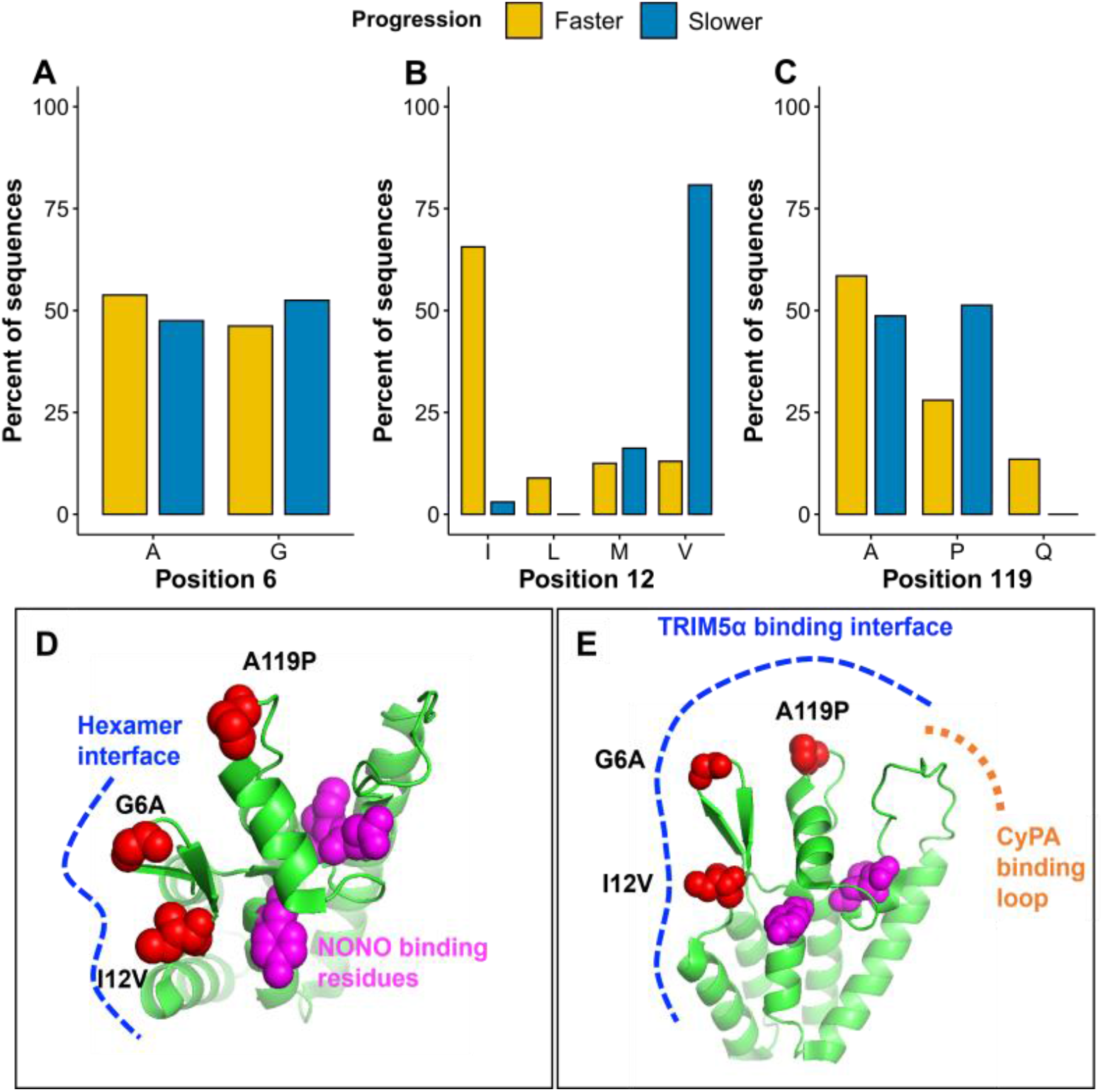
Amino acid sequence diversity in HIV-2 p26 by progression status. **A - C)** The amino acid signature was different at three positions on p26 between faster and slower progressor alignments. The y-axis shows the percentage of sequences from the pooled sequences per progressor group. **D)** This model of HIV-2 p26 NTD (PDB ID: 2WLV) with space-filling models of the three residues (red) and those involved in NONO binding (magenta). The p26 hexamer interface is shown in blue. **E)** The same model as in panel D, but at a different angle, indicating the larger TRIM5α binding interface (blue) and CyPA binding loop (orange).

### Amino acid residues at p26 positions 6, 12 and 119 in the Caio cohort

To test whether the association between p26 amino acids and disease progression held in a larger cohort, we assessed their association with CD4% and HIV-2 viral loads in the Caio cohort. The proportions of amino acids at positions 6, 12 and 119 in the Caio cohort were similar to those in the Police cohort (**Figure S5**). p26 Amino acids were not associated with CD4% in the Caio cohort, and only proline at position 119 was associated with lower HIV-2 plasma viral loads (**Figure S5**).

## Discussion

Synonymous and nonsynonymous evolutionary rates in *p26* were significantly higher in faster disease progressors and correlated negatively with CD4% change rates. This broadly agrees with a previous study of partial HIV-2 *env* from the same cohort which found that faster progressors (determined via the midpoint CD4% or the combined coefficient stratification) had higher evolutionary rates (15). In HIV-1, *p24* synonymous and nonsynonymous evolutionary rates have shown comparable estimates (1.6±1.3×10^−3^ s/s/y and 2.5±4.8×10^−3^ s/s/y, respectively) (45). In our analysis, HIV-2 nonsynonymous evolutionary rates are an order of magnitude lower than the synonymous rates. Findings from a study of interhost evolution of HIV-2 *p26*, showed little evidence of positive selection in HIV-2 *p26* evolution (49). This is surprising as strong Gag-specific CTL responses are common in HIV-2 long-term non-progressors (55). It is possible that the HIV-2 capsid is not able to readily adapt to host immune responses without the virus losing ability to successfully replicate – though this would need further experimental confirmation. If true - this may explain why protection from Gag-specific CTL responses is sustained in HIV-2 long-term non-progressors. This adds further evidence that immune escape, and subsequent positive selection, in the HIV-2 capsid are not associated with disease progression.

Hierarchical phylogenetic model estimates had narrower 95% HPD intervals than the individual relaxed clock models, which is an expected effect of using HPM (56). Besides the larger HPD intervals in the non-HPM estimates, we had good agreement in results between relaxed clock models and the HPM. In addition, hyperprior scales did not significantly affect evolutionary rate estimates, which suggests that our estimates reflected the sequence data and not model parameters’ priors. The preponderance of synonymous substitutions as the main mode of evolution in HIV-2 *p26* suggests that a process which is common to virus replication (either replicative capacity, generation time, or immune activation) are driving both intrahost evolution and disease progression (11,14,57).

Amino acid variation from the MRCA sequence was greater in faster compared with slower progressors. Pairwise diversity and nonsynonymous divergence were also significantly higher in faster progressors. With a sustained higher nonsynonymous evolutionary rate, we would expect a greater accumulation of amino acid diversity in faster progressors compared to slower progressors. Sequence diversity correlates positively with advancing disease progression in HIV-1, and our results suggest a similar finding for HIV-2 (58).

Inferred sites on HIV-2 p26 with structural/functional motifs which have previously been linked to the HIV-1 capsid nucleotide pore, nuclear import of the capsid and NONO binding were completely conserved across all sequences (18–21). This is in line with the conserved nature of retrovirus capsids (21,22,59). At three sites on p26 the most common amino acid differed between faster and slower progressors – 6, 12 and 119. Position 12 flanks a histidine at position 11 which forms part of the nucleotide channel on the capsid (18). The M12I substitution characterised faster progressor sequences, while slower progressors’ most common residue was valine. At position 119 slower progressors often had a proline and faster progressors an alanine. P119 has been associated with lower viral loads and enhanced sensitivity to TRIM5α (23,25,26). However, the latter finding has been contested (60). Position 119 is adjacent to the CyPA binding loop and may interact with TRIM5α. Positions 6 and 12 are located close to the hexamer interface, as well as the CyPA binding loop. This raises the possibility that amino acid variation at these residues can influence the hexamer formation and/or TRIM5α-mediated immune activation. Further investigation is needed, and *in vitro* experiments would be valuable in determining whether any of the variants or, haplotypes thereof, affect virus replicative capacity or HIV-2-specific immune responses. An alternative interpretation of the amino acid diversity near to the p26 hexamer/TRIM5α interface surfaces is that this region has accumulated diversity from ancestral sequences which adapted to primate hosts; and that the mutations identified in this study do not change the HIV-2 capsid’s function in human infection (61,62). In support of this interpretation, we have shown that in a larger cohort, amino acids at positions 6, 12 and 119 were not associated with CD4%, and only A119P was associated with lower HIV-2 plasma viral loads.

Most p26 amino acid variants were stable over time, again indicating that negative selection was dominant in this region’s evolution. We offer two explanations for this: First, the capsid is limited in its adaptive capacity – though this does not explain why we observed significant differences in amino acid sequences between participants. Second, the median time from HIV-2 detection to plasma sample collection was 9.3 years. In *p26* the primary driver of adaptive mutations are CTL responses (63,64). Cytotoxic lymphocyte responses drive HIV adaptation in the early stages after infection, and therefore these changes may not have been captured by our sampling timeframe (65,66).

Limitations to our study include that we analysed a small sample of HIV-2 positive participants (n = 12); however, this is a relatively large number given the logistical difficulties in obtaining HIV-2 RNA from plasma longitudinally. Within this cohort, we were more likely to successfully sequence HIV-2 *p26* in faster progressors which is probably due to the typically higher viral loads in these participants, this also generated a bias in our sample of sequences. Furthermore, all participants were men, who are more likely to have faster disease progression (67,68). Despite this, our results agreed with pre-existing evidence that evolutionary rates correlate with disease progression markers in HIV-2 infection.

Our study highlights what may be a fundamental difference in intrahost evolution between HIV-1 and HIV-2, in that HIV-1 p24 is able to adapt to host immune responses whereas HIV-2 p26 is more limited in this regard (13,69). There are fewer antiretrovirals available to treat HIV-2 than HIV-1; our results indicate that the new generation of direct capsid inhibitors may be attractive options for ART in HIV-2 positive individuals (70,71). HIV-2’s limited ability to diversify the p26 amino acid sequence also suggests that this may be a good target for vaccination strategies.

## Data Availability

All data produced in the present study are available upon reasonable request to the authors

## Additional information

## Acknowledgements

The listed authors and the members of the Sweden Guinea-Bissau Cohort Research (SWEGUB CORE) group, including Babetida N’Buna, Antonio Biague, Ansu Biai, Cidia Camara, Zacarias Jose da Silva, Joakim Esbjörnsson, Marianne Jansson, Sara Karlson, Jacob Lopatko Lindman, Patrik Medstrand, Fredrik Månsson, Hans Norrgren, Angelica A. Palm, Gülsen Özkaya Sahin and Sten Wilhelmson are indebted to the staff of the Police Clinics and the National Public Health Laboratory (LNSP) in Bissau, Guinea-Bissau. We thank Matthew Cotten for providing critical feedback on this manuscript.

## Author contributions

M.T.B., S.R.J., and J.E. conceptualized and designed the study. M.T.B., S.R.J., and J.E. provided funding for the study. The SWEGUB CORE group provided samples from which new sequences used in the study were generated. M.T.B. performed lab work, inferential analyses and produced all figures and tables. J.N. assisted with phylogenetic modelling and data analysis. A.P and S.K processed samples and performed lab work. K.K and K.M performed the structural p26 modelling and interpreted the results. C.O, T.D.S and A.J collected the Caio p26 sequence and clinical data. M.T.B. and J.E. wrote the manuscript and all the authors reviewed, edited, and approved the manuscript for submission.

