## Supplementary materials for "Intrahost evolution of the HIV-2 capsid correlates with progression to AIDS"

#### 1. Participant recruitment

The Police cohort is a prospective open cohort study of police officers recruited from urban and rural areas in Guinea-Bissau. All police officers were invited to join the study and 98% agreed to do so. The cohort started recruiting participants on 6th February 1990 and continued recruitment until 28th September 2009. A total of 4817 people were enrolled and 872 tested HIV positive during the study, 408 HIV-1 positive and 464 HIV-2 positive. HIV-infected participants were followed up until 28th September 2013 and our analysis therefore ends on the 28th of September 2013.

Blood samples for serology and CD4+ T cell counts were collected at enrolment and at scheduled follow-up visits every 12 – 18 months. HIV testing was done at the National Public Health Laboratory in the capital Bissau. HIV disease was classified according to the WHO staging criteria (1). All participants received detailed information about the study before inclusion, as well as pre- and post-test counselling. If a participant died, mortality reports from the Ministry of Interior were crosschecked by health post staff at the main police station in Bissau. Reported symptoms before death were used to classify cause of death. ART was introduced in Guinea-Bissau in 2005 through a national treatment programme. The Police cohort was included in the programme in early 2006. Date of HIV infection was estimated as the mid-time point between the last HIV-seronegative and the first HIV-seropositive sample. Participants with estimated dates of infection are referred to as **seroincident,** and those HIV-2 infected at enrolment as **seroprevalent,** participants.

For this study, participants were drawn from the Guinea-Bissau Police cohort. Twelve HIV-2 infected adults were selected based on availability of plasma samples and meeting the following inclusion criteria:

i) HIV-2 mono-infected

ii) Antiretroviral therapy naïve at the time of plasma sample collection

iii) Longitudinal CD4% data available to classify participants as relative faster or slower disease progressors as described previously

Table S1: Summary epidemiological data for included participants

| Participant | Follow-up time (years) [1] | CD4% time points [2] | Sequence time points [3] |
|---|---|---|---|
| DL2051 | 21.0 | 10 | 3 |
| DL2386 | 18.2 | 4 | 2 |
| DL2524 | 22.6 | 10 | 1 |
| DL2533 | 16.8 | 6 | 1 |
| DL3248 | 4.6 | 3 | 1 |
| DL3286 | 16.8 | 10 | 2 |
| DL3405 | 9.3 | 4 | 2 |
| DL3542 | 20.1 | 9 | 2 |
| DL3646 | 16.1 | 8 | 3 |
| DL3740 | 8.9 | 2 | 3 |
| DL3761 | 9.4 | 2 | 2 |
| DL3941 | 13.3 | 3 | 3 |

[1] Years from HIV-2 detection to last date in the study. [2] Number of CD4% measurements included in the analysis. [3] Number of plasma samples with successful HIV-2 *p26* sequencing.

**Ethics approval and consent to participate**. The study was approved by the ethics committees of the government of Guinea-Bissau; the University of Lund, Sweden; and the Karolinska Institute, Stockholm. Study participants were counselled and provided informed oral consent

#### 2. CD4+ T cell count analysis

Both absolute CD4+ T cell counts and CD4+ T cell percentage (CD4%) are reliable immunological markers of HIV disease progression. In resource-limited settings, CD4% has lower variability and sensitivity to specimen handling, participant age, or time of sampling (2). T-lymphocyte subsets were determined at the National Public Health Laboratory (LNSP), by conventional flow cytometry (Until 2005: FACStrak; Becton Dickinson, San Jose, CA, and from 2006 and onwards: CyFlow, Partec, Münster, Germany). Leucocyte counts were performed with a cell counter until 2005 (Coulter Counter CBC5; Coulter Electronics Ltd, Luton, England), and by CyFlow from 2006 and onwards.

#### 3. Stratification of HIV-2 infected individuals into disease progression groups

Longitudinal follow-up of ART naïve HIV-1 infected individuals has suggested that time to AIDS and death can be explained by CD4+ T cell absolute counts and decline rate as well as viral load (3–6).

For participants with two or more CD4% measurements we analysed per participant linear regression models in R to estimate the midpoint CD4% in follow-up time (**Figure S1**)(7). We used the coefficient of the regression model as the CD4% change rate, measured as CD4% change/year. CD4% increased, stayed the same and decreased for different participants over follow-up; we therefore described CD4% as change rate and not decline rate. We have indicated a negative change rate for a decreasing CD4% with time and a positive change rate for increasing CD4%. This procedure then gave two statistics per participant, a midpoint CD4% level and CD4% change rate. These data were then transformed into proportional values, with each data point expressed as a proportion of the highest value which was 1.0 (highest midpoint CD4% or change rate divided by itself). The transformed data points now fell on a positive scale between 0 and 1. Participants were then ranked from highest CD4% level to lowest, and highest change rate to lowest (they were all on a positive scale at this point). Those with CD4% levels and change rates greater than the mean value were classified as **slower progressors**, those below the mean as **faster progressors**. Participants with their values used in the transformation are shown in **Table S2**.

<u>Table S2: Participant progression stratification</u>

| **Participant** | **Midpoint CD4%** [1] | | **CD4% change rate** [1] | | **Combined coefficient** | **Midpoint CD4%** | **CD4% change rate** | **Combined coefficient** |
|---|---|---|---|---|---|---|---|---|
| DL2051 | 30.7 | 0.9 | -1.8 | 0.4 | 0.66 | Slower | Faster | Faster |
| DL2386 | 25.3 | 0.8 | -1.6 | 0.4 | 0.59 | Faster | Faster | Faster |
| DL2524 | 32.7 | 1.0 | 0.8 | 0.9 | 0.93 | Slower | Slower | Slower |
| DL2533 | 25.1 | 0.8 | 1.1 | 0.9 | 0.84 | Faster | Slower | Slower |
| DL3248 | 29.4 | 0.9 | 1.0 | 0.9 | 0.90 | Slower | Slower | Slower |
| DL3286 | 32.7 | 1.0 | -0.1 | 0.7 | 0.85 | Slower | Slower | Slower |
| DL3405 | 33.1 | 1.0 | 1.5 | 1.0 | 0.99 | Slower | Slower | Slower |
| DL3542 | 30.8 | 0.9 | -1.0 | 0.5 | 0.73 | Slower | Faster | Slower |
| DL3646 | 18.2 | 0.6 | -0.3 | 0.7 | 0.60 | Faster | Slower | Faster |
| DL3740 | 16.5 | 0.5 | 0.7 | 0.8 | 0.67 | Faster | Slower | Faster |
| DL3761 | 16.0 | 0.5 | -3.7 | 0.0 | 0.25 | Faster | Faster | Faster |
| DL3941 | 17.4 | 0.5 | -1.9 | 0.4 | 0.44 | Faster | Faster | Faster |

[1] Midpoint CD4% and CD4% change rates per day are shown in their left columns with the transformed proportional values alongside them. Stratifications into faster and slower progressor groups are shown in the three right-most columns

Finally, to account for variability in disease stages at enrolment we used a combined coefficient derived from an equally weighted estimate using both CD4% level and change rate. To do this, the ranked midpoint CD4% values and CD4% change rates were added together and divided by two. As an example, DL1751's transformed midpoint CD4% value of 0.9 (slower progressor) was added to the transformed CD4% 0.4 (faster progressor) and averaged to give a combined coefficient of 0.66 (faster progressor). This is referred to as the combined coefficient. Participants were finally ranked using this coefficient as faster (above mean value) and slower progressors (below mean value).

#### 4. Viral nucleic acid extraction

Total RNA was extracted from plasma samples which had been stored at -80ºC, using the Qiagen RNeasy Lipid Tissue Mini Kit (Qiagen, Venlo, Netherlands) with minor modifications to the manufacturers' instructions. 100µL of plasma was used for each extraction. 1mL of Qiazol was added to each sample, mixed thoroughly and then incubated at room temperature for 10min. 5µg of carrier RNA was then added followed by 200µL of chloroform. The sample was mixed, then incubated for 3min at room temperature and then centrifuged at 12000xg for 15min at 4ºC in a Megastar 1.6R centrifuge (VWR, Radnor, USA). The aqueous phase was removed and a further 5µg of carrier RNA added to it, followed by an equivalent volume of 70% ethanol. 700µL of the aqueous phase was added to a provided spin column and then centrifuged at 8000xg for 15s at room temperature. The flow through was then removed, and the rest of the sample added to the spin column for centrifugation. After this, 350µL RW1 was added to the sample which was then centrifuged at 8000xg for 15s at room temperature. 10µL of DNase I was added to 70µL of Buffer RDD and mixed. This 80µL solution was then added to the sample and incubated for 15min at room temperature. Next, 350µL RW1 was added, and the sample centrifuged at 8000xg for 15s at room temperature. 500µL of RPE was added and the sample centrifuged at 8000xg for 15s at room temperature. Next a further 500µL RPE was added, and the sample centrifuged at 8000xg for 2min at room temperature RT. The spin column was then placed in a new collection tube, centrifuged at 12000xg for 1min at room temperature. Finally, the spin column was placed in a new 1.5mL tube, 40µL of PCR grade water was added to the membrane and incubated for 10min at room temperature. The sample was then centrifuged at 8000xg for 1min at room temperature.

#### 5. HIV-2 *gag* polymerase chain reaction

**Primer design:** HIV displays significant diversity between participants and also within the quasispecies circulating in a participant at any given time point (8). To design a polymerase chain reaction (PCR) which would amplify the p26 region of *gag* consistently across participants, and across time points, it is necessary to select primers which included conserved sites between participants. We downloaded 356 sequences from the Los Alamos National Laboratory (LANL) sequence database spanning HIV-2 *gag* from position 1103 – 2668 on the HIV-2 BEN.M30502 reference sequence. We then removed short sequences, clonal sequences and all non-group A sequences - the Police cohort is very unlikely to be infected with other HIV-2 groups (9). This left 27 sequences (**Table S3**) which were then aligned in Geneious 2019.2 using the Muscle alignment algorithm (10).

Table S3: HIV-2 *gag* sequences used in primer design

| Number | Accession number [1] |
|---|---|
| 1 | A.GW.1987.CAM2CG.D00835: |
| 2 | A.NL.2003.RH1.MF595865: |
| 3 | A.FR.2000.LA38.KY025539: |
| 4 | A.FR.1998.LA41.KY025542: |
| 5 | A.DE.-.PEI2_KR_KRCG.U22047: |
| 6 | A.JP.2008.NMC786_clone_41.AB731742: |
| 7 | A.DE.-.BEN.M30502: |
| 8 | A.FR.1993.LA37.KY025538: |
| 9 | A.PT.-.ALI.AF082339: |
| 10 | A.IN.2007.NNVA.EU980602: |
| 11 | A.FR.1996.LA40.KY025541: |
| 12 | A.CI.1988.UC2.U38293: |
| 13 | A.FR.2001.LA42.KY025543: |
| 14 | A.FR.2002.LA36GomM.KU168287: |
| 15 | A.FR.1998.LA39.KY025540: |
| 16 | A.IN.1995.CRIK_147.DQ307022: |
| 17 | A.GH.-.GH1.M30895: |
| 18 | A.GW.1986.FG_clone_NIHZ.J03654: |
| 19 | A.GW.-.MDS.Z48731: |
| 20 | A.GM.1987.D194.X52223: |
| 21 | A.GM.-.MCR35.AY509260: |
| 22 | A.SN.1986.ST_JSP4_27.M31113: |
| 23 | A.SN.1985.ROD.M15390: |
| 24 | A.GM.-.ISY_SBL_6669_85.J04498: |
| 25 | A.GM.-.MCN13.AY509259: |
| 26 | A.GW.1987.CAM2.KU179861: |
| 27 | A.IN.-.NIM_8.DQ973520: |

[1] Accession numbers for sequences on the LANL database.

We designed primers for a nested PCR to flank the p26 region of HIV-2 BEN.M30502. Eight forward and six reverse degenerate primers were selected and tested in a variety of primer pairs (**Figure S1**). We selected one pair of outer primers and one pair of inner primers (**Table S4**). The nested PCR was able to amplify p26 consistently at an HIV-2 RNA concentration of 200 copies/mL. Below this level amplification became inconsistent. All PCRs were run on a LifeTouch thermal cycler (Bioer, Technology, Tokyo, Japan)

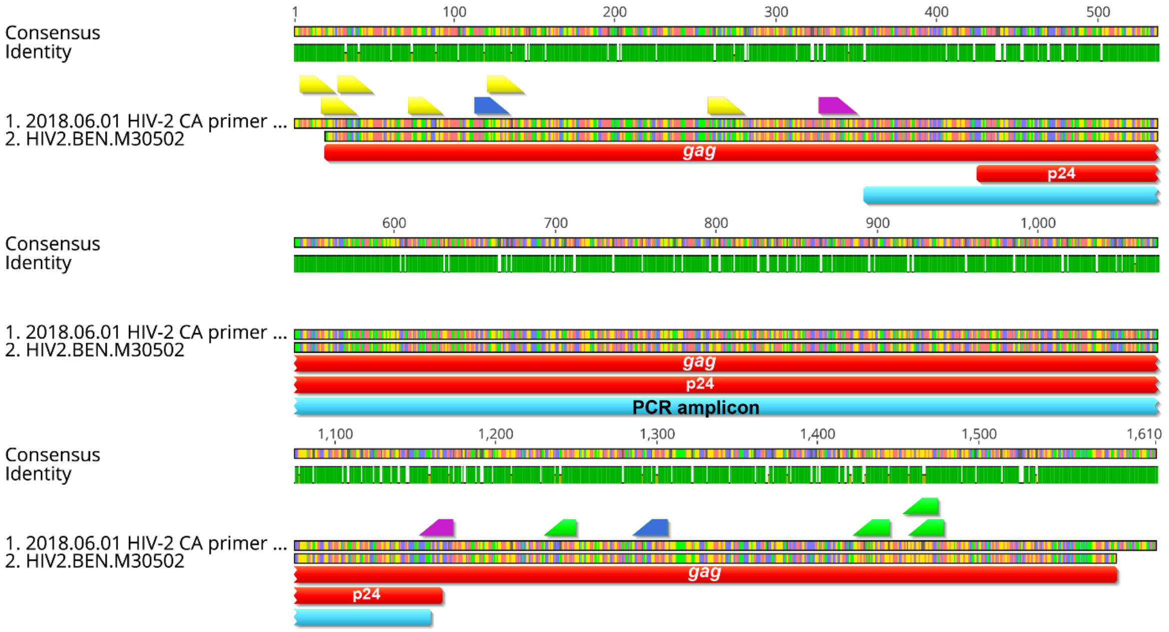


**Figure S1: Primer design for HIV-2 *gag* PCR**. The HIV-2 *gag* sequences alignment consensus sequence (2018.06.01 HIV-2 CA primer) is shown mapped to HIV-2 BEN.M30502. Identity agreement at the nucleotide level is shown with green bar indicating 100% agreement. The consensus of this pairwise alignment is also shown. Forward primers are highlighted in yellow, reverse in green. Outer primers used in the nested reaction are shown in blue, inner primers in purple. Annotations for the maximum inner amplicon length and the p26 region are shown.

Table S4: Primers used in the HIV-2 *gag* PCR

| **Primers** | **Oligo sequence** | **Binding site** [1] | **Amplicon size** [2] |
|---|---|---|---|
| Outer primers | | | |
| HIV2CA_IF4 | AAACATRTTGTGTGGGCAGCGA | 1197 - 1218 | 1193 |
| HIV2CA_IR2 | CTGTCTWTCTGGGCARTTTGCC | 2369 - 2390 | |
| Inner primers | | | |
| HIV2CA_IF2 | TAGTACAGAGACATCTAGYGGCAG | 1411 - 1434 | 846 |
| HIV2CA_IR3 | TGYTGGGCTGCTGCRAATGGG | 2237 - 2257 | |

[1] Binding site in reference to HIV-2 BEN.M30502 from 5' to 3'. [2] Amplicon length shown here includes the primer sequences. Forward primers are shown 5' to 3' and reverse primers are shown 3' to 5'. Degenerate primers were selected to allow amplification on sequences with variable bases.

Nested PCRs include two stages of amplification. The first step involved a one-step reverse transcription PCR (**RT-PCR**) using the SuperScript IV One-Step RT-PCR System with Platinum Taq DNA Polymerase (Thermo Fisher Scientific, Waltham, MA) (**Table S5**). RT-PCR's start with reverse transcription: the RT-PCR master mix includes an RT enzyme. 5µL of extracted RNA was added to 15.5µL of master mix which was then incubated at 50ºC for 30min followed by the PCR with the outer primers (**Table S6**). The outer set of primers amplified a 1193 nucleotide base pair (bp) fragment from nucleotide 1197 – 2390 of HIV-2 BEN.M30502 (**Figure S1**). After the RT-PCR 2µL of the amplified outer product was used as a template for the **nested reaction**. The nested reaction with the inner set of primers was performed using the Dream Taq PCR kit (Thermo Fisher). A negative control was carried through from RNA extraction to RT-PCR and the final nested reaction. The final sequenced PCR product represented a single open reading frame 735 nucleotides long and mapped to nucleotide 1460 – 2195 on HIV-2 BEN.M30502. PCR products were purified using the in-house EXO-SAP protocol and stored at -20ºC until further downstream applications.

**Exo-Sap master mix per reaction:**

**Reagents:** Recombinant exonuclease I (New England Biolabs, Ipswich, USA) - 0.1µL per reaction, Recombinant shrimp alkaline phosphatase (New England Biolabs) - 0.5µL and PCR grade H20 - 0.4µL. 1.0µL of the EXO-SAP mix was then added to each PCR product and incubated on a thermocycler as follows: 1 cycle 37 °C for 45min and 1 cycle 80 °C for 25min

<u>Table S5: Reagents per reaction for HIV-2 *gag* PCR</u>

| **RT-PCR** | **Nested reaction** |
|---|---|
| 12.5µL 2X buffer | 16.875µL PCR-grade $H_{2}0$ |
| 1.0µL forward primer (100ng/L) | 2.5µL 10X Dream Taq buffer |
| 1.0µL reverse primer (100ng/L) | 2.5µL dNTP (2mM) |
| 1.0µL enzyme mix (RT + Taq) | 1µL forward primer (100ng/L) |
| - | 1µL reverse primer (100ng/L) |
| - | 0.125µL Dream Taq DNA polymerase (5U/L) |

Table S6: Cycling conditions for HIV-2 *gag* PCR

| PCR step | RT-PCR | Nested PCR |
|---|---|---|
| 1. Incubation | 50 °C 30 min | - |
| 2. Denaturation | 94ºC - 2min | 94ºC - 2min |
| 3.1 Denaturation | 94ºC - 15s | 94ºC - 30s |
| 3.2 Annealing | 53ºC - 15s | 55ºC - 30s |
| 3.3 Elongation | 68ºC - 60s | 72ºC - 90s |
| 4. Final elongation | 68ºC - 5min | 72ºC - 5min |

#### 6. Cloning and sequencing

Twenty-five separate PCR products were amplified from the 12 participants, nine of the participants successfully amplified multiple PCR products from different time points and three had only a single time point successfully amplified. The PCR products were cloned using a TOPO-TA cloning kit (Invitrogen, Carlsbad, CA, USA) (**Figure S2**).

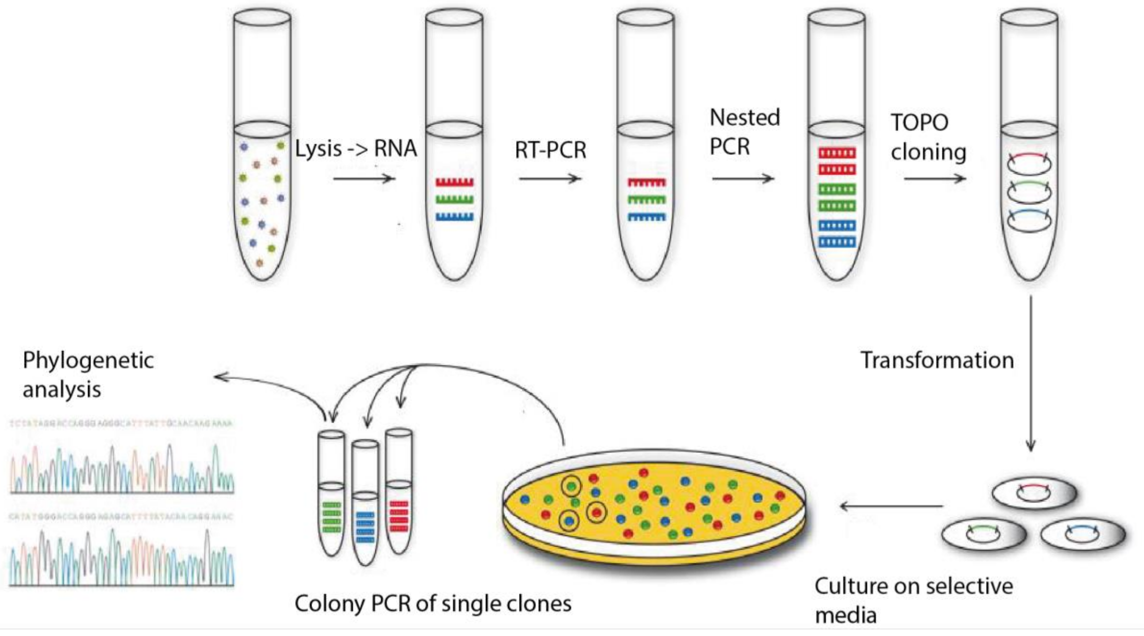


**Figure S2: Cloning process for HIV-2 *gag* final PCR products**. Steps involved in cloning and DNA sequencing of HIV-2 *gag* amplicons.

TOPO-TA cloning was performed in three separate steps: ligation, transformation, and colony PCR.

**Ligation:** 4µL of purified HIV-2 *gag* PCR product was aliquoted into a clean 1.5mL Eppendorf tube. 1µL of salt solution and 1µL of pCRTM4-TOPO plasmid stock was added and gently mixed by tapping the tube. This was then incubated for 30min at room temperature.

**Transformation:** For each vector (PCR product ligated to plasmid) one vial of chemically competent E. coli TOP10 cells was thawed from -80ºC on ice. 5µL of vector was added to one vial, gently mixed and incubated on ice for 30min. The cells were then heat-shocked at 42ºC for 30s in a water bath. Cells were then rested on ice for 2min. 250µL of sterile Super Optimal broth with Catabolite repression Media was added to each vial. Cells were then incubated at 37ºC for 1h in a shaking incubator rotating at 225rpm. After incubation 75µL of each vial was added to kanamycin (50µg/mL) selective Luria-Bertani media growth plates and incubated for 16h at 37ºC. Vector only negative controls were added to each experiment to exclude contamination between plates.

**Colony PCR:** 23 colonies were picked randomly and amplified in a colony PCR using the Advantage 2 PCR kit (Takara). The colony PCR used the same primers as the nested reaction described above and gave an amplicon of identical length to the insert. The colony PCR products were then sequenced with the inner amplification primers with Macrogen Europe (Amsterdam, Netherlands) using Sanger sequencing. Clonal sequences from each time point represent the circulating quasispecies.

### 7. Phylogenetic sequence analysis

#### HIV-2 *p26* sequence cladogram

A total of 369 heterochronous sequences were included in the final analysis. No clear clustering was noted by progression status, and within host sequences from participants formed monophyletic clusters with high posterior support values.

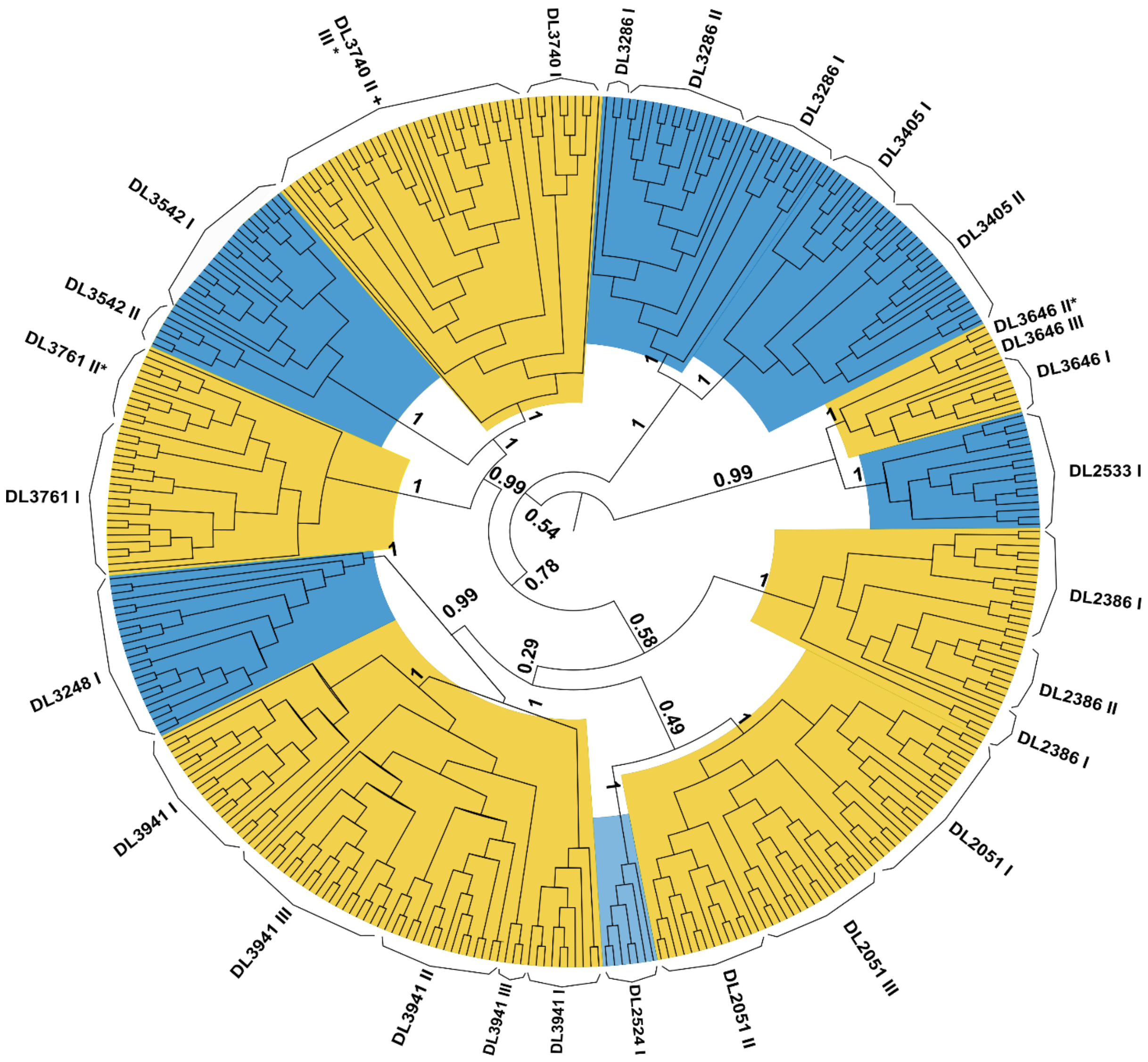


**Figure S3: Cladogram of all HIV-2 *p26* sequences included in phylogenetic analysis.** This maximum clade credibility (MCC) cladogram represents the average of the posterior tree samples generated in the MRCA reconstruction. There is clear monophyletic clustering of participant-specific sequences with high posterior support values of 1.0. Faster disease progressors are highlighted in yellow and slower progressors in blue. An asterisk indicates that participant taxa were mixed by time point.

Table S7: Accession numbers for sequences used in HIV-2 subtyping analysis:

| HIV/SIV | Group | Accession number |
|---|---|---|
| HIV-2 | A | A.GW.2010.TD062.MH681609 |
| HIV-2 | A | A.IN.1995.CRIK_147.DQ307022 |
| HIV-2 | A | A.CI.1988.UC2.U38293 |
| HIV-2 | A | A.PT.-.ALI.AF082339 |
| HIV-2 | A | A.DE.-.BEN.M30502 |
| HIV-2 | A | A.DE.-.PEI2_KR_KRCG.U22047 |
| HIV-2 | A | A.NL.2001.RH2.7.MF595863 |
| HIV-2 | B | B.CI.-.IC763088.AF073842 |
| HIV-2 | B | B.CI.-.IC3983.AF073841 |
| HIV-2 | B | B.CI.-.IC2380.AF073840 |
| HIV-2 | B | B.CI.-.IC763129.AF073843 |
| HIV-2 | B | B.JP.2001.IMCJ_KR020_1_.AB100245 |
| HIV-2 | B | B.CI.-.20_57.AB485671 |
| HIV-2 | B | B.CI.-.20_56.AB485670 |
| HIV-2 | C | L33077 |
| HIV-2 | D | L33083 |
| HIV-2 | E | L33087 |
| HIV-2 | F | F.SL.1993.93SL2F.U75441 |
| HIV-2 | F | KP890355 |
| HIV-2 | G | AF208027 |
| HIV-1 | A | 02A1.GH.1997.97GH-AG2.AB052867 |
| HIV-1 | A1 | A1.UG.1992.UG029.AB098332 |
| HIV-1 | A1 | A1.UG.1992.UG029.AB098333 |
| HIV-1 | AE | 01_AE.TH.1995.95TNIH022.AB032740 |
| HIV-1 | AE | 01_AE.TH.1995.95TNIH047.AB032741 |
| HIV-1 | AE | 01_AE.JP.1993.93JP_NH1.AB052995 |
| HIV-1 | AE | 01_AE.JP.1993.NH25_93JPNH25T_93JP_NH2_5T.AB070352 |
| HIV-1 | AG | 02_AG.GH.1997.97GH-AG1.AB049811 |
| HIV-1 | B | B.FR.1983.LAI-J19.A07867 |
| HIV-1 | B | B.JP.-.PARTICIPANT_IMS1.AB074049 |
| HIV-1 | B | B.JP.-.PARTICIPANT_IMS1.AB074050 |
| HIV-1 | C | C.IN.1993.93IN101.AB023804 |
| SIVsmm | SIVsmm | SMM.US.-.RM174.V2.tf.KU182923 |
| SIVsmm | SIVsmm | SMM.US.-.FTq.tf.KU182920 |
| SIVsmm | SIVsmm | SMM.US.2011.SIVsmE660-FL8.JQ864086 |
| SIVsmm | SIVsmm | SMM.US.-.SME543.U72748 |

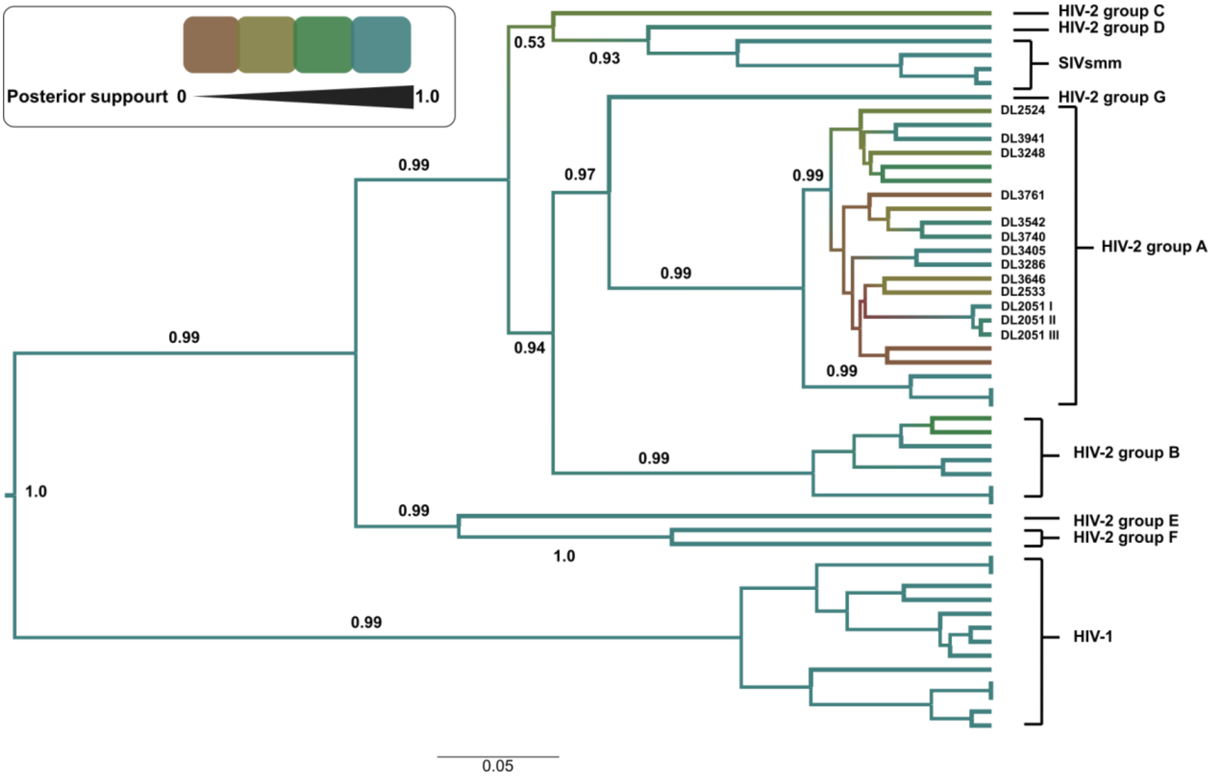


**Figure S4: Phylogenetic tree of subtyping analysis for HIV-2 *gag* sequences.** The MCC shows posterior support values for all internal branches with the colour scale shown in the top left corner. High support values of 0.99 indicate that the cloned *gag* sequences were HIV-2 group A. Scale bar shows genetic distance from the tree root.

#### HIV-2 capsid molecular clock estimates

All relaxed clock models were run using the same parameters as described in the methods section. Only participants with multiple time points (n = 9) were included. The HPM model tested a series of fixed effects as explanatory variables for the evolutionary rates, under a strict molecular clock prior.

<u>Table S8: Evolutionary rate summaries for relaxed and strict clock models</u>

| Participant | **Relaxed clock model evolutionary rates** | | **HPM strict clock model evolutionary rates** | | Progression status |
|---|---|---|---|---|---|
| | median | 95% HPD | median | 95% HPD | |
| DL2051 | $5.4x10^{-3}$ | $2.8x10^{-3}$ - $8.6x10^{-3}$ | $3.5x10^{-3}$ | 2.5 - $5.7x10^{-3}$ | faster |
| DL2386 | $4.0x10^{-3}$ | $6.4x10^{-4}$ - $1.2x10^{-2}$ | $2.8x10^{-3}$ | 1.9 - $3.7x10^{-3}$ | faster |
| DL3286 | $1.1x10^{-3}$ | $4.7x10^{-4}$ - $2.0x10^{-3}$ | $1.4x10^{-3}$ | 5.8 - $2.3x10^{-3}$ | slower |
| DL3405 | $3.6x10^{-3}$ | $1.3x10^{-3}$ - $6.4x10^{-3}$ | $2.1x10^{-3}$ | 1.0 - $4.5x10^{-3}$ | slower |
| DL3542 | $2.7x10^{-3}$ | $1.4x10^{-3}$ - $4.3x10^{-3}$ | $2.9x10^{-3}$ | 1.8 - $3.7x10^{-3}$ | slower |
| DL3646 | $3.9x10^{-3}$ | $1.9x10^{-3}$ - $7.3x10^{-3}$ | $3.1x10^{-3}$ | 2.2 - $4.0x10^{-3}$ | faster |
| DL3740 | $3.1x10^{-3}$ | $9.1x10^{-4}$ - $9.3x10^{-3}$ | $3.1x10^{-3}$ | 1.1 - $4.7x10^{-3}$ | faster |
| DL3761 | $1.5x10^{-2}$ | $1.6x10^{-3}$ - $4.7x10^{-2}$ | $3.7x10^{-3}$ | 2.4 - $8.9x10^{-3}$ | faster |
| DL3941 | $1.1x10^{-2}$ | $4.18x10^{-3}$ - $2.0x10^{-2}$ | $3.3x10^{-3}$ | 2.4 - $5.0x10^{-3}$ | faster |

Participant relaxed clock model estimates for molecular clock rates with their 95% highest posterior density are shown. Progressor status as determined by the combined coefficient. Participant HPM estimates for molecular clock rates with their 95% highest posterior density (HPD) are shown.

#### 8. Association between HIV-2 p26 amino acids, CD4% and HIV-2 viral load in the Caio cohort

We analysed the effect of amino acids at position 6, 12 and 119 on CD4% and HIV-2 viral load in the Caio cohort (n = 86) (11,12). CD4% measurements were available for 59/86 participants, with eight having one measurement and 51 more than one. The mean CD4% was calculated for each participant and compared by p26 amino acid residue, an approach that would be equivalent with estimated midpoint CD4% in follow up. HIV-2 plasma viral loads were available for 60/86 patients (eight with one measurement and 52 with multiple), and mean log10 viral loads were compared by p26 amino acid.

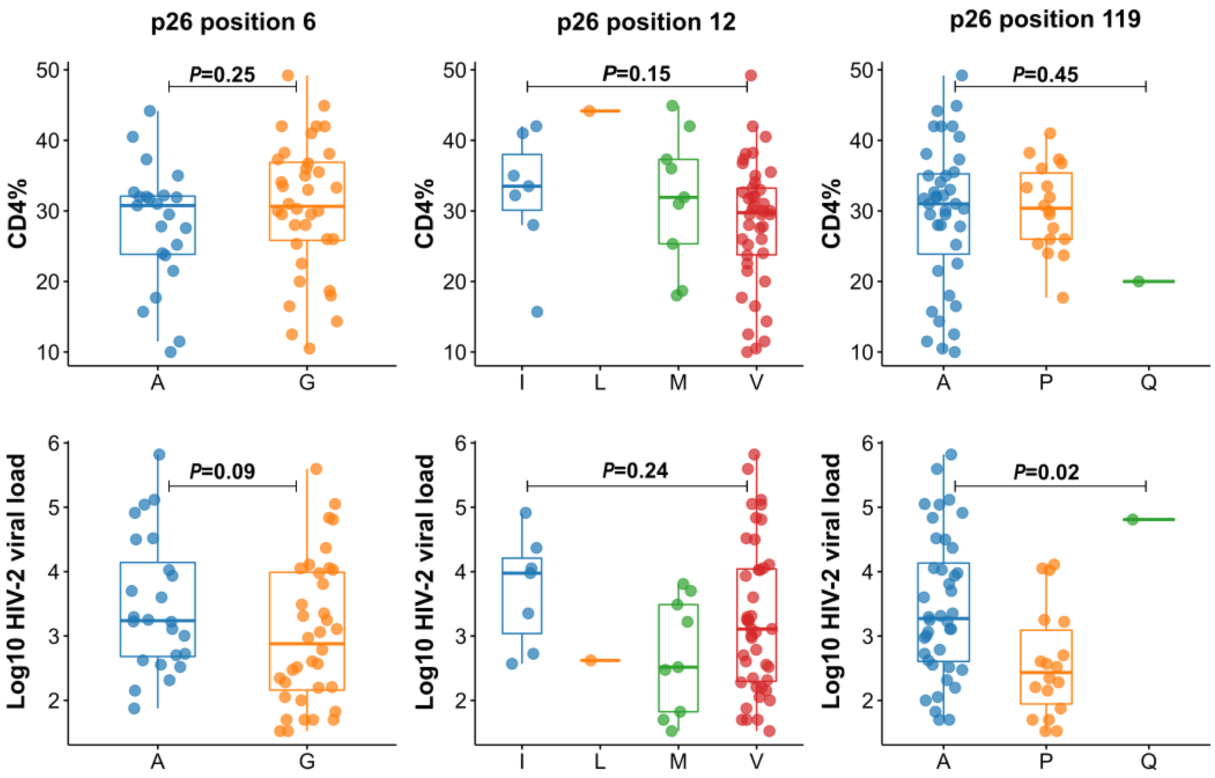


**Figure S5: CD4% and HIV-2 plasma viral load by amino acid at p26 positions 6, 12 and 119.** Top panels: amino acids at positions 6, 12 and 119 were not associated with significant differences in CD4% after correction for multiple comparisons. Bottom panels: Proline at position 119 was associated with lower HIV-2 plasma viral loads (log10 HIV-2 viral load: A119 = 3.3 [IQR: 2.6-4.1] vs 119P = 2.4 [IQR:1.9-3.1], MW *P*=0.03 after correction for multiple comparisons done using False Discovery Rate). *P* values for Kruskall-Wallis tests are shown.

#### References


1. WHO case definitions of HIV for surveillance and revised clinical staging and immunological classification of HIV-related disease in adults and children HIV/AIDS Programme. 2007.
